# Heterogeneous treatment effects of intensive glycemic control on kidney microvascular outcomes in ACCORD

**DOI:** 10.1101/2023.06.14.23291396

**Authors:** Vivek Charu, Jane W. Liang, Glenn M. Chertow, Zhuo Jun Li, Maria E. Montez-Rath, Pascal Geldsetzer, Ian H. de Boer, Lu Tian, Manjula Kurella Tamura

## Abstract

**Objective:** Clear criteria to individualize glycemic targets are lacking. In this post-hoc analysis of the Action to Control Cardiovascular Risk in Diabetes trial (ACCORD), we evaluate whether the kidney failure risk equation (KFRE) can identify patients who disproportionately benefit from intensive glycemic control on kidney microvascular outcomes.

**Research design and methods:** We divided the ACCORD trial population in quartiles based on 5-year kidney failure risk using the KFRE. We estimated conditional treatment effects within each quartile and compared them to the average treatment effect in the trial. The treatment effects of interest were the 7-year restricted-mean-survival-time (RMST) differences between intensive and standard glycemic control arms on (1) time-to-first development of severely elevated albuminuria or kidney failure and (2) all-cause mortality.

**Results:** We found evidence that the effect of intensive glycemic control on kidney microvascular outcomes and all-cause mortality varies with baseline risk of kidney failure. Patients with elevated baseline risk of kidney failure benefitted the most from intensive glycemic control on kidney microvascular outcomes (7-year RMST difference of 115 v. 48 days in the entire trial population) However, this same patient group also experienced shorter times to death (7-year RMST difference of -57 v. -24 days).

**Conclusions:** We found evidence of heterogenous treatment effects of intensive glycemic control on kidney microvascular outcomes in ACCORD as a function of predicted baseline risk of kidney failure. Patients with higher kidney failure risk experienced the most pronounced benefits of treatment on kidney microvascular outcomes but also experienced the highest risk of all-cause mortality.

## Introduction

Type 2 diabetes is a widespread non-communicable disease of increasing prevalence wherein complications contribute to morbidity and mortality, requiring staggering healthcare expenditures^1^. The leading cause of kidney failure, blindness, peripheral neuropathy, and lower limb amputation, type 2 diabetes is also a dominant risk factor for atherosclerotic cardiovascular disease and mortality. Effective glycemic control is fundamental to diabetes management; for patients who require more than non-pharmacological (lifestyle) management strategies, an array of oral and injectable medications have been introduced over the past two decades. Despite an expansion of therapeutic options, there remains uncertainty regarding the benefits and risks of the intensity of glycemic control for patients with type 2 diabetes^2,3^.

Previously conducted randomized controlled trials have shown that intensive glycemic control reduces microvascular events and may reduce macrovascular events, but increases the risk of serious hypoglycemic events and may increase the risk of mortality^4–7^. Although newer pharmacological therapies have reduced the risks of serious hypoglycemia, nearly all clinical diabetes management guidelines advocate personalizing glycemic targets to reduce the potential for harm^8–12^. Unfortunately, clear criteria for how to individualize glycemic targets are lacking^13^. Intensive glycemic control has consistently been shown to reduce the risk of microvascular disease, explained largely by its effect on albuminuria^14^. Therefore, identifying patients who differentially benefit from intensive glycemic control on kidney microvascular outcomes would be a promising approach to individualize glycemic targets.

Heterogeneous treatment effects occur when the effect of treatment depends on the baseline covariates. Conventional subgroup analyses (e.g. testing “one-variable-at-a-time” interactions) in RCTs of intensive glycemic control have failed to identify meaningful heterogeneity in treatment effects on microvascular outcomes^14^. These analyses, however, have only explored heterogeneous treatment effects on the hazard ratio/relative scale, despite the fact that such effects are best understood on the absolute scale^15^. Indeed, the lack of heterogeneity of treatment effects on the relative scale does not imply lack of heterogeneity of treatment effects on the absolute scale. Perhaps more importantly, conventional approaches to subgroup analysis are underpowered, prone to spurious false positive results due to multiple testing, and under-represent true clinical heterogeneity, in which patients differ from one another across many variables simultaneously^16^. Instead, modern predictive approaches to characterizing heterogeneous treatment effects group patients using many clinically salient variables simultaneously, and are better at detecting important treatment effect heterogeneity obscured by conventional analyses^15–18^.

In this work, we employed a risk-based approach to identifying heterogeneous treatment effects, in which a multivariable model that predicts risk for the outcome is used to stratify patients within the trial to quantify risk-based variation in treatment effects. In this post-hoc analysis of ACCORD, we evaluated whether the kidney failure risk equation (KFRE), a validated tool that integrates patient’s baseline age, sex, estimated glomerular filtration rate, and urine albumin-to-creatinine ratio into a risk score for kidney failure, identified patients who could disproportionately benefit from intensive glycemic control^19,20^.

## Methods

### Data

This is a post-hoc secondary analysis of the limited-access ACCORD BioLINCC dataset obtained from the United States National Institutes of Health. Details of the ACCORD study population, interventions and study procedures have been previously published^4,7^. In brief, 10251 patients with diabetes, HbA1c greater than 7.5% and cardiovascular disease (two or more cardiovascular risk factors), were randomly assigned to intensive glycemic control (HbA1c<6.0%) or standard glycemic control (HbA1c 7.0 to 7.9%).

### Outcomes

The primary outcomes of interest in this analysis were: (1) kidney microvascular events, defined as the composite of the time to first development of severely elevated albuminuria (urine albumin-to-creatinine ratio >=300 mg/g) or time to first development of kidney failure, defined as initiation of maintenance dialysis or kidney transplantation, or an increase in serum creatinine > 3.3 mg/dL in the absence of an acute reversible cause; (2) all-cause mortality. Kidney microvascular events were pre-specified outcomes in ACCORD, and the original definitions are used here^7^.

### Treatment effect of interest

There are several metrics used to define treatment effects with time-to-event data. Heterogeneous treatment effects are best understood on the absolute risk scale, and as such, the treatment effect of interest here is the 7-year restricted mean survival time (RMST) difference for each of the outcomes under intensive versus standard glycemic control^17,21,22^. The RMST captures the average time free from a clinical event, within a specific time window (hence, “restricted”). The RMST difference, a measure of the treatment effect, is the average time delay in the onset of an event under treatment versus control, within a specific time window. It has been shown that one can make inferences about the RMST up to the largest follow-up time in the study^23^. As such, in this analysis, we focus on the 7-year RMST difference between intensive and standard glycemic control, as in the ACCORD study, patients were followed for up to 7 years. The RMST difference has an additional geometric interpretation as the area between the two estimated survival curves of interest.

### Statistical analysis

We analyzed the data in accordance with the intention-to-treat principle, with the goal of quantifying heterogeneous treatment effects of intensive glycemic control on kidney microvascular outcomes and all-cause mortality. Heterogeneous treatment effects occur when the effect of treatment varies with baseline covariates in a non-random way. In this work, we employ the “risk-modeling” approach, in which a multivariable model that predicts risk for the outcome is applied to stratify patients within the trial to examine risk-based variation in treatment effects. The premise behind using risk-modeling to identify heterogeneous treatment effects is that the effect of treatment will vary with baseline risk of the outcome (also called “risk magnification”)^15,17,18^. While this is not universally true, several successful applications of risk-modeling to identify heterogenous treatment effects have been published in large cardiovascular trials. Risk-based analyses using multivariable risk prediction tools are better powered than conventional subgroup analyses, and have a lower risk of false-positive findings^16^. Though a risk-model can be developed using data from the RCT itself, an externally developed prediction model is preferred since over-fitting data in the trial population can exaggerate the degree of risk heterogeneity. Externally developed prediction models are also more likely to be generalizable and have clinical utility. As such, in this work, we utilized externally developed risk prediction models to explore heterogenous treatment effects of intensive glycemic control on kidney outcomes^19,20,24^. If treatment effects truly varied with patients’ baseline risk of the outcome, we would expect a biologically plausible relationship between risk and the treatment effect (e.g. a monotone increasing treatment effect with risk or a U-shaped curve between risk and treatment effects).

For participants with available data, we calculated the 5-year risk of kidney failure using the kidney failure risk equation (KFRE), which incorporates information on baseline age, sex, estimated glomerular filtration rate (using the race-free CKD-EPI 2021 equation), and urine albumin-to-creatinine ratio. We estimated the empirical distribution of 5-year risk of kidney failure in the entire trial population, and determined the 25th, 50th and 75th percentiles of the empirical distribution. We grouped patients into quartiles of their 5-year risk of kidney failure.

Using the overall trial population, we estimated the average treatment effect as the 7-year RMST difference between intensive and standard glycemic control for the composite kidney microvascular outcome and all-cause mortality (average treatment effect). Randomization is not guaranteed to achieve covariate balance within subgroups. Therefore, within each quartile of predicted 5-year risk of kidney failure, before estimating treatment effects, we first quantified the balance of the baseline covariates across the treatment and control arms. Baseline variables with absolute standardized mean differences >0.10 would require adjustment in the RMST estimation, e.g. by using an ANCOVA-type adjustment for these covariates. Within each quartile, we estimated the 7-year RMST difference between intensive and standard glycemic control for the composite kidney microvascular outcome and all-cause mortality (conditional average treatment effects).

We quantified evidence for heterogeneous treatment effects by calculating the difference between the average treatment effect in the entire trial population and the conditional average treatment effects within each subgroup defined by quartiles of predicted 5-year risk of kidney failure. We estimated standard errors and associated 95% confidence intervals for the conditional average treatment effects as well as their differences with the average treatment effect via a bootstrap procedure with 1000 replicates. A significant non-zero difference (associated 95% CI does not contain zero) between the average treatment effect and the conditional average treatment effect in a subgroup indicates evidence of treatment effect heterogeneity.

To allow for comparison with conventional metrics of treatment effects in time-to-event data, we also present estimated hazard ratios using Cox’s proportional hazards models.

### Companion analyses

In companion analyses, we explored treatment effects within the quartiles of 5-year predicted kidney failure risk on the time to first medically-attended hypoglycemic event, a relevant adverse event in the setting of glycemic control. We also explored treatment effects within deciles of 5-year predicted kidney failure risk. Finally, we implemented an alternative risk model that specifically predicts early diabetic kidney disease^24^. As with the KFRE, we evaluated for heterogeneous treatment effects within quartiles of predicted diabetic kidney disease risk based on the alternative model.

### Reproducibility

All code to reproduce this analysis is available at https://github.com/janewliang/ACCORD_KFRE.

## Results

Of 10251 participants randomized in ACCORD, 9777 (4904 randomized to intensive and 4873 randomized to standard glycemic control; 95.3%) had available baseline covariate data (age, sex, estimated glomerular filtration rate, and urine albumin-to-creatinine ratio) to estimate 5-year predicted kidney failure risk via the Kidney Failure Risk Equation (KFRE). The average effect of intensive glycemic control in the trial eligible population was to delay the onset of severely elevated albuminuria or kidney failure by 48.4 days over a 7-year period (7-year RMST difference: 48.4 days [95% CI: 25.3 - 69.6]; corresponding hazard ratio: 0.75 [95% CI: 0.65– 0.86]; **Figure S1**). In contrast, the average effect of intensive glycemic control was to shorten the time to death by 23.6 days over a 7-year period (7-year RMST difference: -23.6 days [95% CI: - 42.2 to -6.6]; hazard ratio: 1.20 [95% CI: 1.04–1.40]; **Figure S1**).

Our goal was to assess whether effects of intensive glycemic control on kidney microvascular outcomes and all-cause death varied with patients’ baseline risk of kidney failure. The distributions of 5-year predicted kidney failure risk based on the KFRE were similar in the intensive and standard glycemic control arms (**Figure S2**). The 25th, 50th and 75th percentiles of the empirical distribution of 5-year predicted kidney failure risk in the entire trial population were used to divide trial participants into four mutually exclusive subgroups. The 5-year predicted kidney failure risk ranged between zero and 0.004% (mean: 0.002%) in the first quartile; 0.004 and 0.014% (mean: 0.008%) in the second; 0.014 and 0.078% (mean: 0.034%) in the third; and 0.078 and 97% (mean: 0.99%) in the fourth. **Table 1** displays the baseline characteristics of patients in each of the four subgroups. Compared to patients with the lowest predicted 5-year risk of kidney failure (quartile 1 [Q1]), those with the highest risk of kidney failure (quartile 4 [Q4]) were more likely to be male, Black, and older in age, have lower fasting plasma glucose, higher urine albumin-to-creatinine ratio, higher serum creatinine and lower eGFR (**Table 1**). We found that baseline covariates were well-balanced between the treatment and control arms within each subgroup; absolute standardized mean differences were uniformly below 0.10 (**Figure S3**).

**Table 1.**
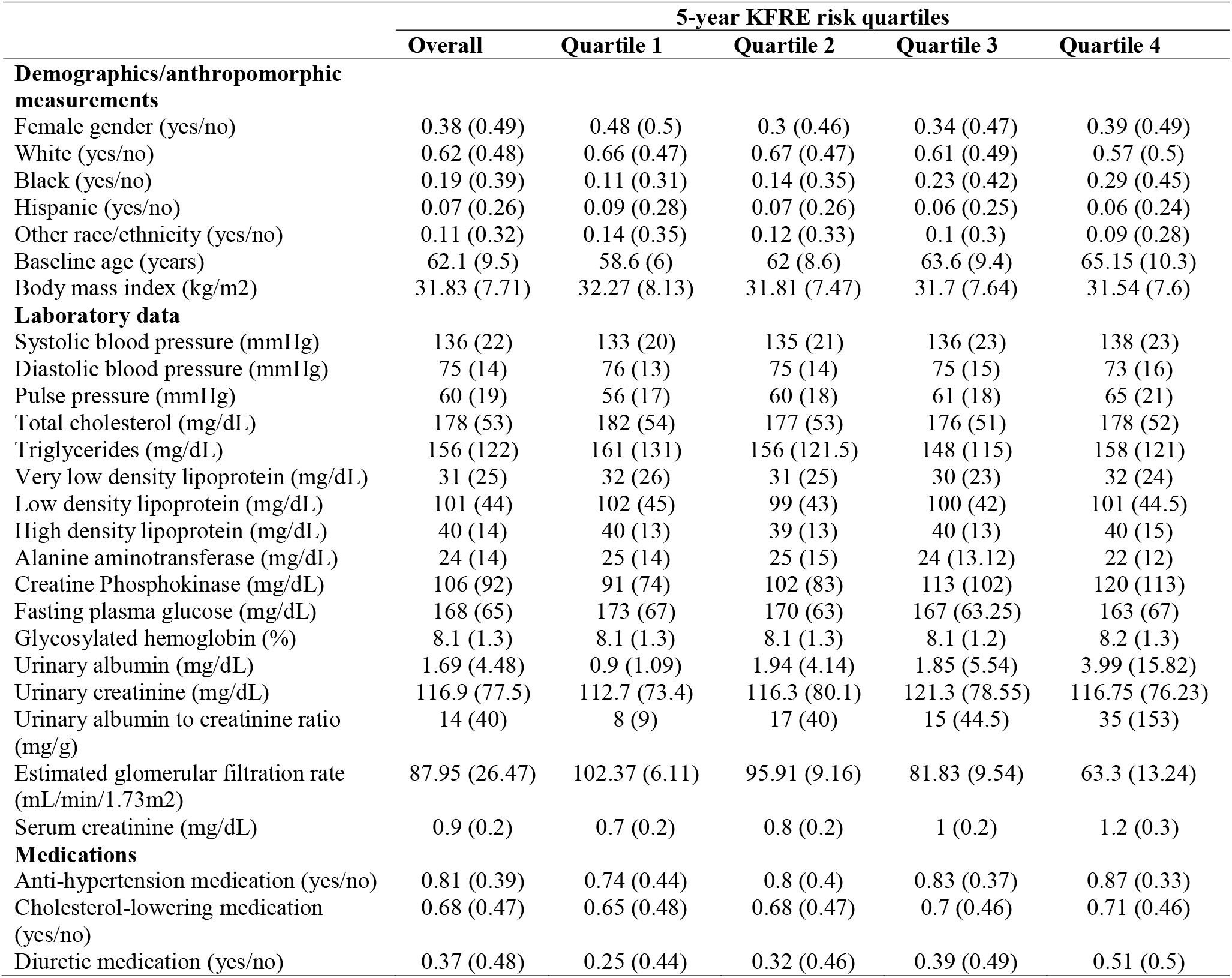
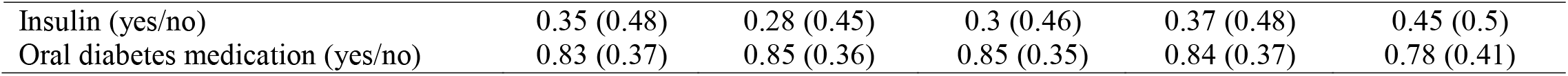
Baseline characteristics of ACCORD patients, overall, and by quartile of 5-year predicted risk of kidney failure via the kidney failure risk equation. Data are presented as mean (SD).

We found evidence that the effect of intensive glycemic control on kidney microvascular outcomes varied with baseline risk of kidney failure (**Figure 1; Figure 2**). First, we quantified how the effect of intensive glycemic control within each of the four subgroups differs from the overall average treatment effect in the trial. In the absence of heterogenous treatment effects, the effect of intensive glycemic control in each subgroup would be equivalent to the average treatment effect in the entire trial population, and thus the difference in treatment effects between each subgroup and the overall trial population would be zero. In contrast, we found that compared to the average treatment effect in the entire trial population, patients in Q1 had lower treatment effects (difference in RMST differences between the overall trial effect and Q1 effect: −38.3 days [95% CI: −71.0 to −6.3]), and patients in Q4 experienced more than double the average treatment effect (difference in RMST differences between the overall trial effect and Q4 effect: 66.4 days [95% CI: 19.5 to 118.1]; **Figure 1, Table 2)**. Patients in Q2 and Q3 experienced treatment effects similar to the average treatment effect (**Figure 1**). More specifically, translating these findings to the original treatment effect scale, we estimate that for patients with 5-year predicted kidney failure risk less than 0.004% (Q1), intensive glycemic control delayed the onset of severely elevated albuminuria or kidney failure by 10.1 days over a 7-year window (95% CI: −23.9 to 43.1), while for patients with a 5-year predicted kidney failure risk greater than 0.08% (Q4), intensive glycemic control delayed this onset by 114.8 days (95% CI: 58.1 to 176.4; **Table 2**). These results indicate that the average treatment effect observed in the entire trial population was predominantly driven by the effect seen in patients with elevated 5-year kidney failure risk. In companion analyses, we further demonstrated that the effect of intensive glycemic control on the composite kidney outcome in patients in the highest quartile of kidney failure risk is largely driven by its effect on the incident severely elevated albuminuria outcome (**Figure S4**). We also demonstrated consistent, monotone, results when considering deciles of predicted 5-year kidney failure risk (rather than quartiles; **Figure S5**).

**Table 2.** Heterogeneous treatment effects of intensive glycemic control on the composite kidney outcome and all-cause death. Treatment effects in bold have 95% CI that do not overlap with zero. *The normalized restricted mean survival time (RMST) difference in days, defined as the RMST difference in the subgroup of interest minus the RMST difference in the entire trial. Normalized RMST values of zero indicate that the treatment effect in the subgroup of interest is equivalent to that in the entire trial population (no evidence of heterogeneous treatment effects). Normalized RMST values above zero (including the 95% confidence interval) indicate that the treatment effect in the subgroup of interest is larger than the treatment effect in the entire trial population (more beneficial); values below zero indicate that the treatment effect in the subgroup of interest is below that in the entire trial population (more harmful). Abbreviations: RMST = restricted mean survival time; Q1-Q4 = quartiles 1-4

| Population/outcome of interest | 7-year RMST difference (95% CI; days) | Difference between the 7-year RMST difference in each quartile and that in the overall trial* (95% CI; days) | Hazard Ratio (95% CI) |
| --- | --- | --- | --- |
| **Composite kidney outcome** |  |  |  |
| Overall trial | **48.40 (25.34, 69.60)** | Ref. | **0.75 (0.65-0.86)** |
| KFRE Q1 | 10.14 (-23.9, 43.13) | **-38.25 (-70.97, -6.26)** | 0.93 (0.63-1.37) |
| KFRE Q2 | 40.07 (-4.65, 84.7) | -8.33 (-46.88, 31.06) | 0.78 (0.59-1.04) |
| KFRE Q3 | **50.08 (7.95, 92.69)** | 1.68 (-35.00, 36.96) | **0.73 (0.54-0.83)** |
| KFRE Q4 | **114.77 (58.14, 176.41)** | **66.37 (19.51, 118.09)** | **0.66 (0.53-0.83)** |
| **All-cause mortality** |  |  |  |
| Overall trial | **-23.64 (-42.2, -6.64)** | - | **1.20 (1.04-1.40)** |
| KFRE Q1 | -21.03 (-47.7, 6.66) | 2.61 (-21.95, 30.27) | 1.37 (0.91-2.07) |
| KFRE Q2 | -19.53 (-53.51, 12.56) | 4.11 (-27.23, 32.29) | 1.15 (0.84-1.58) |
| KFRE Q3 | 11.06 (-20.23, 42.14) | **34.7 (7.29, 63.81)** | 0.95 (0.70-1.29) |
| KFRE Q4 | **-56.7 (-100.19, -17.50)** | **-33.06 (-66.43, -0.53)** | **1.30 (1.03-1.64)** |

**Figure 1.**
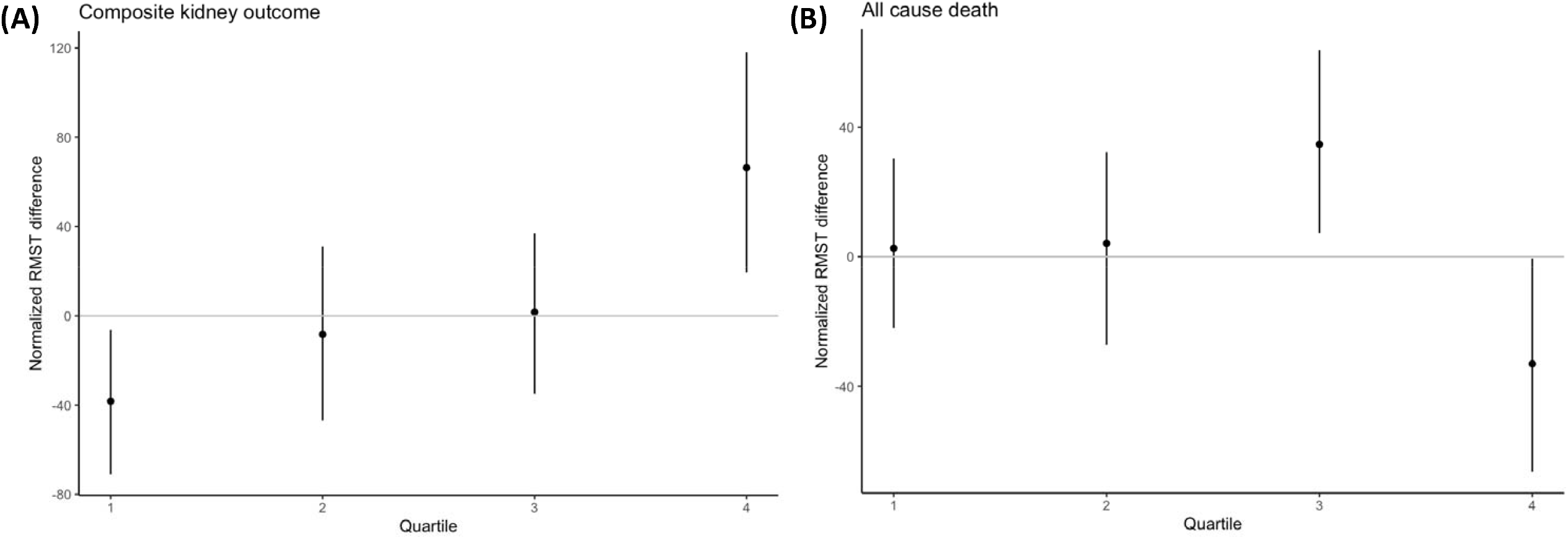
Heterogeneous treatment effects of intensive glycemic control on (A) the composite kidney outcome and (B) all-cause death. The x-axis displays each subgroup of patients, defined by quartiles of 5-year predicted risk by the KFRE. The y-axis displays the normalized restricted mean survival time (RMST) difference in days, defined as the RMST difference in the subgroup of interest minus the RMST difference in the entire trial. Normalized RMST values of zero indicate that the treatment effect in the subgroup of interest is equivalent to that in the entire trial population (no evidence of heterogeneous treatment effects). Normalized RMST values above zero (including the 95% confidence interval) indicate that the treatment effect in the subgroup of interest is larger than the treatment effect in the entire trial population (more beneficial); values below zero indicate that the treatment effect in the subgroup of interest is below that in the entire trial population (more harmful).

**Figure 2.**
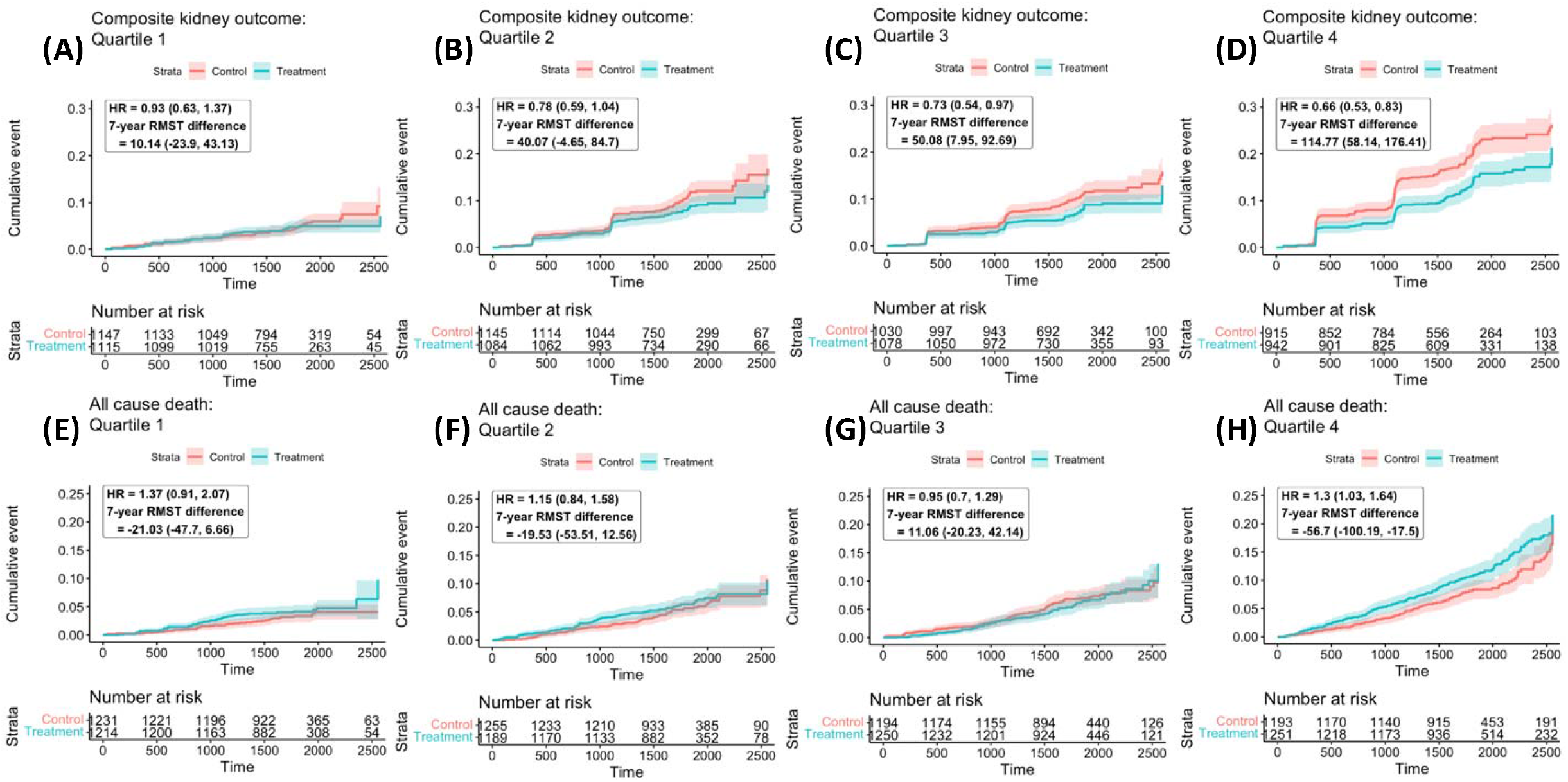
(A-D): Heterogeneous treatment effects of intensive versus standard glycemic control on the composite kidney outcome, by quartile of KFRE 5-year predicted risk at baseline. Plots demonstrate cumulative incidence curves by quartile of KFRE 5-year predicted risk at baseline; hazard ratios and the 7-year restricted mean survival time difference between treatment and control arms are presented in days. (E-H) Heterogeneous treatment effects of intensive versus standard glycemic control on all-cause mortality, by quartile of KFRE 5-year predicted risk at baseline. Plots demonstrate cumulative incidence curves by quartile of KFRE 5-year predicted risk at baseline; hazard ratios and the 7-year restricted mean survival time difference between treatment and control arms are presented in days.

We also found evidence that the effect of intensive glycemic control on all-cause mortality varies with baseline risk of kidney failure (**Figure 4B; Figure 6**). The effect of intensive glycemic control on all-cause mortality for patients in Q1 and Q2 of 5-year predicted kidney failure risk were similar to the overall population; however, patients in Q3 experienced a lower risk of mortality than average (difference in RMST differences between the overall trial effect and Q3 effect: 34.7 days [95% CI: 7.3 to 63.8]; **Figure 4B**) and patients in Q4 experienced a higher risk of mortality than average (difference in RMST differences between the overall trial effect and Q4 effect: -33.1 days [95% CI: -66.4 to -0.5]; **Figure 4B**). Translating these comparisons to the original treatment effect scale, we estimate that for patients in Q4 of 5-year predicted kidney failure risk, intensive glycemic control shortens the time to death by 56.7 days over a 7-year time frame (7-year RMST difference: -56.7 days [95% CI: -100.2 to -17.5]).

In companion analyses, we found evidence that the effect of intensive glycemic control on the time-to-first-attended hypoglycemic event also varied with baseline KFRE risk; patients with the highest risk of kidney failure experienced the shortest time-to-first-hypoglycemic event on intensive glycemic control (**Figure S6-S7**). We assessed how an alternative risk score for diabetic kidney disease performed in identifying heterogeneous treatment effects; we found no evidence for heterogeneous treatment effects of intensive glycemic control on the composite kidney outcome or all-cause mortality when grouping patients based on the diabetic kidney disease risk score developed by Jiang et al^24^. (**Figures S8-S10**).

## Discussion

In this post-hoc analysis of ACCORD, we demonstrated heterogeneous absolute treatment effects of intensive glycemic control (target HgbA1C <6.0%) on kidney microvascular outcomes and all-cause mortality based on baseline 5-year predicted risk of kidney failure using the kidney failure risk equation (KFRE). We found that patients in the highest quartile of 5-year risk of kidney failure at baseline (greater than 0.08% by KFRE) benefitted disproportionately from intensive glycemic control on the composite kidney outcome (incident severely elevated albuminuria or kidney failure; 7-year RMST difference of 114.8 days [95% CI: 58.1 to 176.4]), but also experienced an increased risk of all-cause death (7-year RMST difference: −56.7 days [95% CI: −100.2 to −17.5]). Absolute treatment effects in the highest quartile of kidney failure risk were approximately two-to-three-fold those in the entire trial population.

There remains debate about the role of intensive glycemic control in the management of patients with type II diabetes. While nearly all guidelines recommend individualized glycemic targets, the American Diabetes Association (ADA), International Diabetes Federation (IDF) and European Association for the Study of Diabetes (EASD) recommend an A1C target of less than 7% for most adults with type II diabetes; the American Association for Clinical Endocrinologists (AACE) recommends an HgbA1C target of less than 6.5%. Based on the results of several large clinical trials, including ACCORD, the primary benefit of intensive glycemic control is reduced risk of microvascular complications, mostly driven by kidney microvascular outcomes^14^. Our analysis demonstrates that the observed effect of intensive glycemic control on renal microvascular outcomes in ACCORD is almost entirely driven by a subset of patients representing one-quarter of the trial eligible population at elevated risk of kidney failure at baseline. To our knowledge, ours is the first study to demonstrate that the benefit of intensive glycemic control on renal microvascular outcomes was most pronounced in a subset of patients in ACCORD.

Several studies have explored potential mechanisms for the increase in mortality observed among participants randomized to intensive glycemic control in ACCORD^25–29^. Our finding that patients at higher risk of kidney failure at baseline also experienced higher risks of mortality with intensive glycemic control echoes prior work demonstrating increased cardiovascular mortality under intensive glycemic control for patients with CKD in ACCORD^30^. Definitive explanations to account for the increased mortality in this group of patients, however, are lacking. Importantly, other large randomized trials of intensive glycemic control, ADVANCE and VADT, did not demonstrate increased mortality among patients receiving intensive glycemic control, and, in the modern era, new pharmacologic strategies may allow for glycemic control with lower risks of hypoglycemia than with insulin and/or sulfonylurea agents used in ACCORD. Whether our findings in ACCORD are reproducible in other randomized trials of intensive glycemic control, VADT and ADVANCE, remains to be seen^5,6^.

Our study has several strengths. First, we quantify treatment effects on the absolute scale, using the RMST metric. The RMST captures the mean survival time in each treatment arm over a time window, with the difference in RMSTs describing the delay in onset of the outcome of interest between the treatment arms. In contrast to the hazard ratio, the RMST has a causal interpretation and a direct interpretation^31–33^ Second, we employed a risk-modeling approach to identify heterogenous treatment effects of intensive glycemic control on renal microvascular outcomes, utilizing an externally-validated risk score. The risk-modeling approach aims to subgroup patients based on baseline risk of the outcome using multiple clinically meaningful baseline covariates, avoiding potential issues with conventional “one-variable-at-a-time” subgroup identification. While this approach has been successfully employed to characterize heterogeneous treatment effects in several cardiovascular trials^18^, it has been underutilized in kidney disease trials. To our knowledge, this is the first study exploring heterogenous treatment effects on renal outcomes in ACCORD, and the first study to evaluate whether treatment effects on kidney outcomes might vary with a well-validated risk score for kidney failure. Additionally, our study highlights several critical methodological considerations when exploring heterogenous treatment effects in randomized trials using similar approaches: (1) it is essential to consider treatment effects on both potential benefits and adverse events – our analysis demonstrates the subgroup that derived the largest absolute benefit on kidney outcomes from intensive glycemic control also experienced the largest absolute risk for hypoglycemic events and all-cause mortality; (2) the choice of risk score is essential in studies employing a risk-modeling approach – we were not able to identify meaningful heterogenous treatment effects using an alternative risk score developed for diabetic kidney disease (see supplement)^24^. Overall, our approach is straightforward, and could have promising applications to several newer kidney disease clinical trials, including those of sodium-glucose transport protein 2-inhibitors and glucagon-like-1 receptor agonists. To encourage applications of these methods, we have made code publicly available.

Several limitations must also be mentioned. First, we used the KFRE to estimate patients’ 5-year risk of kidney failure at baseline^19,20^. The KFRE was developed to predict kidney failure in patients with CKD stages 3-5, while the majority of ACCORD participants did not have CKD at baseline; we note that the KFRE is often used in patients with early-stage CKD (stage 2) as well^34,35^. We did also assess heterogenous treatment effects using an alternative risk score that specifically predicts early diabetic kidney disease, but we failed to identify meaningful heterogeneity in treatment effects on renal outcomes using this score^24^. It is possible that an alternative risk score would provide a more optimal grouping of patients than the KFRE as well. Second, while risk-modeling is an elegant approach to identifying heterogenous treatment effects, the underlying assumption of the approach is that treatment effects will vary with baseline risk for the outcome. While this is certainly reasonable in many clinical contexts, it is not guaranteed, mathematically or clinically. Lastly, kidney function was only evaluated at scheduled visits causing interval censoring and the precise time of severely elevated albuminuria or kidney failure was unknown in general, which may reduce the power of the analysis. However, due to randomization, it is unlikely that interval censoring introduced systematic bias in estimating the treatment effects.

A validated tool to predict kidney failure identified individuals who gain the largest absolute kidney benefit, but also experienced the largest absolute risk of death from the ACCORD intensive glycemic control treatments. Our findings illustrate the value of applying modern predictive approaches to uncover clinically meaningful treatment heterogeneity for diabetes and kidney disease therapies, particularly when treatments have effects on multiple endpoints.

## Data Availability

This is a post-hoc secondary analysis of the limited-access ACCORD BioLINCC dataset obtained from the United States National Institutes of Health. Code to reproduce the analysis presented is available at: https://github.com/janewliang/ACCORD_KFRE

## Guarantor

VC takes full responsibility for the work as a whole, including the study design, access to data, and the decision to submit and publish the manuscript.

## References

1. Khan MAB, Hashim MJ, King JK, Govender RD, Mustafa H, Al Kaabi J. Epidemiology of Type 2 Diabetes – Global Burden of Disease and Forecasted Trends. J Epidemiol Glob Health. 2020;10(1):107–111. doi:10.2991/jegh.k.191028.001

2. Rodriguez-Gutierrez R, Gonzalez-Gonzalez JG, Zuñiga-Hernandez JA, McCoy RG. Benefits and harms of intensive glycemic control in patients with type 2 diabetes. BMJ. 2019;367:l5887. doi:10.1136/bmj.l5887

3. Rodríguez-Gutiérrez R, Montori VM. Glycemic Control for Patients With Type 2 Diabetes Mellitus: Our Evolving Faith in the Face of Evidence. Circulation: Cardiovascular Quality and Outcomes. 2016;9(5):504–512. doi:10.1161/CIRCOUTCOMES.116.002901

4. Action to Control Cardiovascular Risk in Diabetes Study Group, Gerstein HC, Miller ME, et al. Effects of intensive glucose lowering in type 2 diabetes. N Engl J Med. 2008;358(24):2545–2559. doi:10.1056/NEJMoa0802743

5. ADVANCE Collaborative Group, Patel A, MacMahon S, et al. Intensive blood glucose control and vascular outcomes in patients with type 2 diabetes. N Engl J Med. 2008;358(24):2560–2572. doi:10.1056/NEJMoa0802987

6. Duckworth W, Abraira C, Moritz T, et al. Glucose control and vascular complications in veterans with type 2 diabetes. N Engl J Med. 2009;360(2):129–139. doi:10.1056/NEJMoa0808431

7. Ismail-Beigi F, Craven T, Banerji MA, et al. Effect of intensive treatment of hyperglycaemia on microvascular outcomes in type 2 diabetes: an analysis of the ACCORD randomised trial. Lancet. 2010;376(9739):419–430. doi:10.1016/S0140-6736(10)60576-4

8. American Diabetes Association. 6. Glycemic Targets: Standards of Medical Care in Diabetes—2021. Diabetes Care. 2020;44(Supplement_1):S73–S84. doi:10.2337/dc21-S006

9. de Boer IH, Caramori ML, Chan JCN, et al. Executive summary of the 2020 KDIGO Diabetes Management in CKD Guideline: evidence-based advances in monitoring and treatment. Kidney International. 2020;98(4):839–848. doi:10.1016/j.kint.2020.06.024

10. Blonde L, Umpierrez GE, Reddy SS, et al. American Association of Clinical Endocrinology Clinical Practice Guideline: Developing a Diabetes Mellitus Comprehensive Care Plan—2022 Update. Endocrine Practice. 2022;28(10):923–1049. doi:10.1016/j.eprac.2022.08.002

11. de Boer IH, Khunti K, Sadusky T, et al. Diabetes Management in Chronic Kidney Disease: A Consensus Report by the American Diabetes Association (ADA) and Kidney Disease: Improving Global Outcomes (KDIGO). Diabetes Care. 2022;45(12):3075–3090. doi:10.2337/dci22-0027

12. Kidney Disease: Improving Global Outcomes (KDIGO) Diabetes Work Group. KDIGO 2022 Clinical Practice Guideline for Diabetes Management in Chronic Kidney Disease. Kidney Int. 2022;102(5S):S1–S127. doi:10.1016/j.kint.2022.06.008

13. Ismail-Beigi F, Moghissi E, Tiktin M, Hirsch IB, Inzucchi SE, Genuth S. Individualizing Glycemic Targets in Type 2 Diabetes Mellitus: Implications of Recent Clinical Trials. Ann Intern Med. 2011;154(8):554–559. doi:10.7326/0003-4819-154-8-201104190-00007

14. Zoungas S, Arima H, Gerstein HC, et al. Effects of intensive glucose control on microvascular outcomes in patients with type 2 diabetes: a meta-analysis of individual participant data from randomised controlled trials. Lancet Diabetes Endocrinol. 2017;5(6):431–437. doi:10.1016/S2213-8587(17)30104-3

15. Kent DM, Paulus JK, van Klaveren D, et al. The Predictive Approaches to Treatment effect Heterogeneity (PATH) Statement. Ann Intern Med. 2020;172(1):35–45. doi:10.7326/M18-3667

16. Kent DM, Rothwell PM, Ioannidis JP, Altman DG, Hayward RA. Assessing and reporting heterogeneity in treatment effects in clinical trials: a proposal. Trials. 2010;11(1):85. doi:10.1186/1745-6215-11-85

17. Kent DM, van Klaveren D, Paulus JK, et al. The Predictive Approaches to Treatment effect Heterogeneity (PATH) Statement: Explanation and Elaboration. Ann Intern Med. 2020;172(1):W1–W25. doi:10.7326/M18-3668

18. Kent DM, Steyerberg E, Klaveren D van. Personalized evidence based medicine: predictive approaches to heterogeneous treatment effects. BMJ. 2018;363:k4245. doi:10.1136/bmj.k4245

19. Tangri N, Stevens LA, Griffith J, et al. A Predictive Model for Progression of Chronic Kidney Disease to Kidney Failure. JAMA. 2011;305(15):1553–1559. doi:10.1001/jama.2011.451

20. Tangri N, Grams ME, Levey AS, et al. Multinational Assessment of Accuracy of Equations for Predicting Risk of Kidney Failure: A Meta-analysis. JAMA. 2016;315(2):164–174. doi:10.1001/jama.2015.18202

21. Chen PY, Tsiatis AA. Causal inference on the difference of the restricted mean lifetime between two groups. Biometrics. 2001;57(4):1030–1038. doi:10.1111/j.0006-341x.2001.01030.x

22. Kloecker DE, Davies MJ, Khunti K, Zaccardi F. Uses and Limitations of the Restricted Mean Survival Time: Illustrative Examples From Cardiovascular Outcomes and Mortality Trials in Type 2 Diabetes. Ann Intern Med. 2020;172(8):541–552. doi:10.7326/M19-3286

23. Tian L, Jin H, Uno H, et al. On the empirical choice of the time window for restricted mean survival time. Biometrics. 2020;76(4):1157–1166. doi:10.1111/biom.13237

24. Jiang W, Wang J, Shen X, et al. Establishment and Validation of a Risk Prediction Model for Early Diabetic Kidney Disease Based on a Systematic Review and Meta-Analysis of 20 Cohorts. Diabetes Care. 2020;43(4):925–933. doi:10.2337/dc19-1897

25. Basu S, Raghavan S, Wexler DJ, Berkowitz SA. Characteristics Associated With Decreased or Increased Mortality Risk From Glycemic Therapy Among Patients With Type 2 Diabetes and High Cardiovascular Risk: Machine Learning Analysis of the ACCORD Trial. Diabetes Care. 2018;41(3):604–612. doi:10.2337/dc17-2252

26. Bonds DE, Miller ME, Bergenstal RM, et al. The association between symptomatic, severe hypoglycaemia and mortality in type 2 diabetes: retrospective epidemiological analysis of the ACCORD study. BMJ. 2010;340:b4909. doi:10.1136/bmj.b4909

27. Boyko EJ. ACCORD Glycemia Results Continue to Puzzle. Diabetes Care. 2010;33(5):1149–1150. doi:10.2337/dc10-0432

28. Riddle MC, Ambrosius WT, Brillon DJ, et al. Epidemiologic relationships between A1C and all-cause mortality during a median 3.4-year follow-up of glycemic treatment in the ACCORD trial. Diabetes Care. 2010;33(5):983–990. doi:10.2337/dc09-1278

29. Siraj ES, Rubin DJ, Riddle MC, et al. Insulin Dose and Cardiovascular Mortality in the ACCORD Trial. Diabetes Care. 2015;38(11):2000–2008. doi:10.2337/dc15-0598

30. Papademetriou V, Lovato L, Doumas M, et al. Chronic kidney disease and intensive glycemic control increase cardiovascular risk in patients with type 2 diabetes. Kidney International. 2015;87(3):649–659. doi:10.1038/ki.2014.296

31. Hernán MA. The Hazards of Hazard Ratios. Epidemiology. 2010;21(1):13–15. doi:10.1097/EDE.0b013e3181c1ea43

32. Martinussen T, Vansteelandt S, Andersen PK. Subtleties in the interpretation of hazard contrasts. Lifetime Data Anal. 2020;26(4):833–855. doi:10.1007/s10985-020-09501-5

33. Royston P, Parmar MK. Restricted mean survival time: an alternative to the hazard ratio for the design and analysis of randomized trials with a time-to-event outcome. BMC Medical Research Methodology. 2013;13(1):152. doi:10.1186/1471-2288-13-152

34. Duggal V, Montez-Rath ME, Thomas IC, Goldstein MK, Tamura MK. Nephrology Referral Based on Laboratory Values, Kidney Failure Risk, or Both: A Study Using Veterans Affairs Health System Data. Am J Kidney Dis. 2022;79(3):347–353. doi:10.1053/j.ajkd.2021.06.028

35. Bhachu HK, Cockwell P, Subramanian A, et al. Impact of Using Risk-Based Stratification on Referral of Patients With Chronic Kidney Disease From Primary Care to Specialist Care in the United Kingdom. Kidney Int Rep. 2021;6(8):2189–2199. doi:10.1016/j.ekir.2021.05.031

