## Supplementary Materials for "Heterogeneous treatment effects of intensive glycemic control on kidney microvascular outcomes in ACCORD"

**Figure S1.** Effects of intensive (treatment) versus standard (control) glycemic control in the ACCORD trial population on (A) the composite kidney outcome and (B) all-cause mortality. 7-year RMST differences are presented in days.

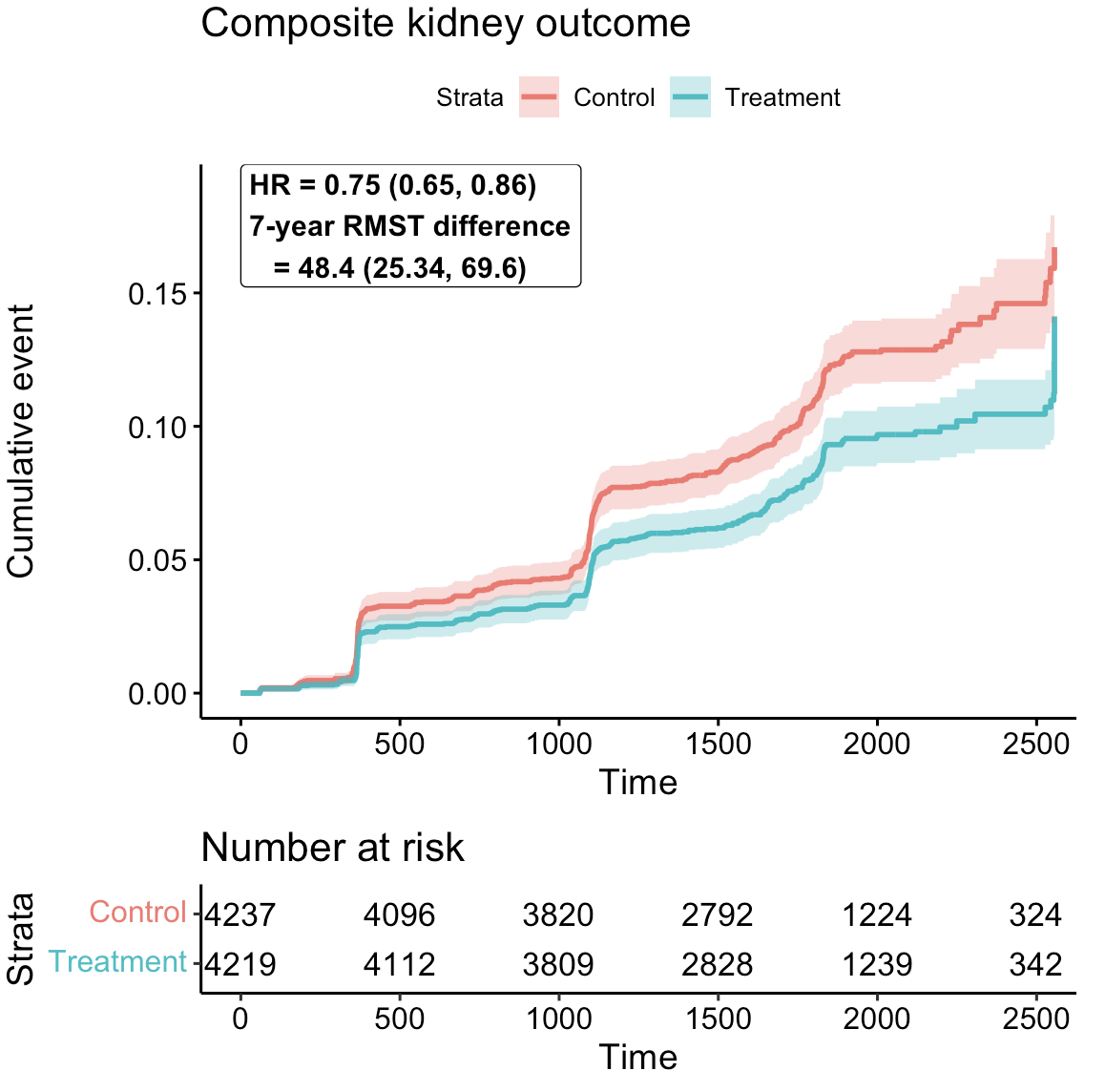

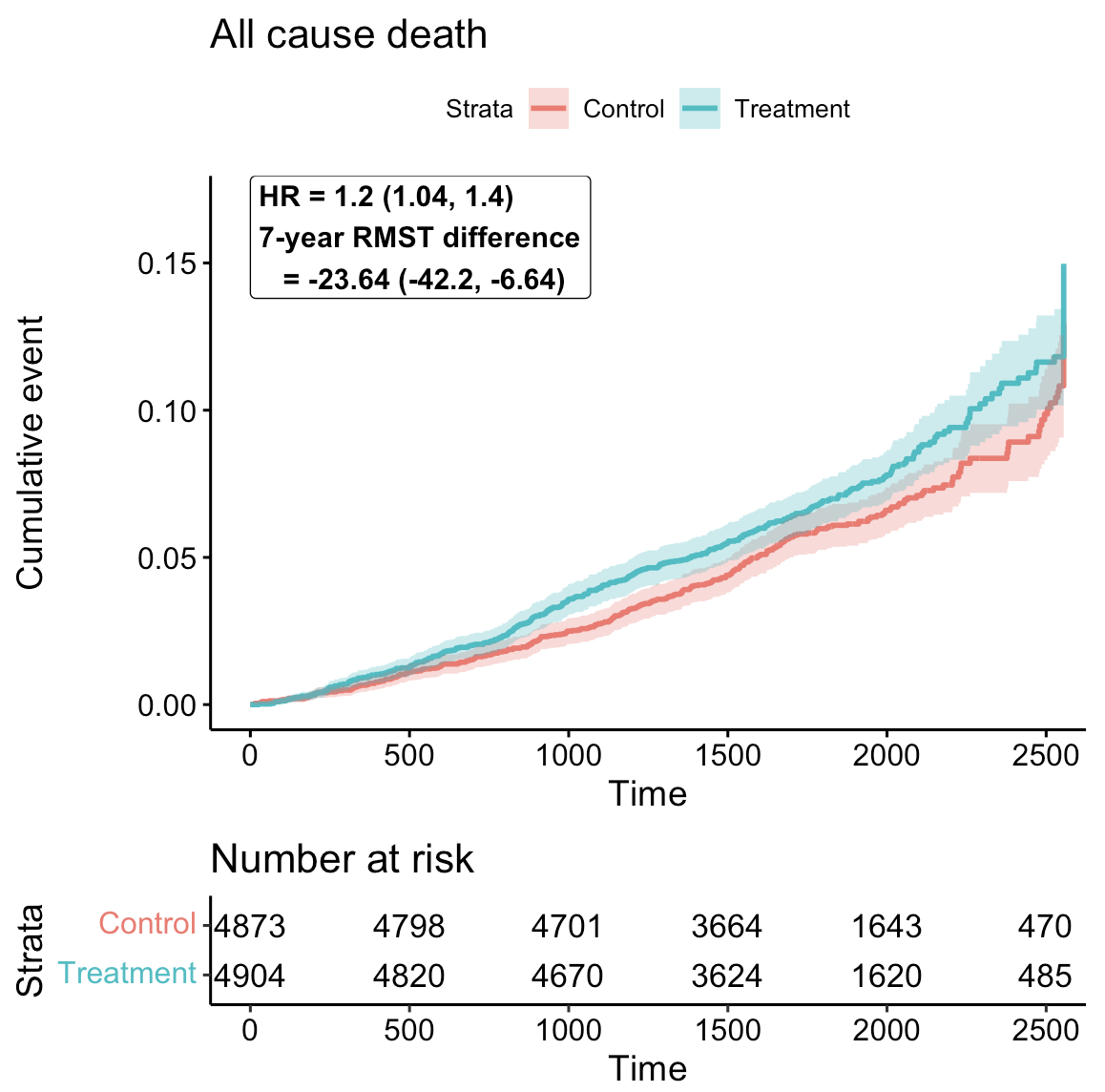

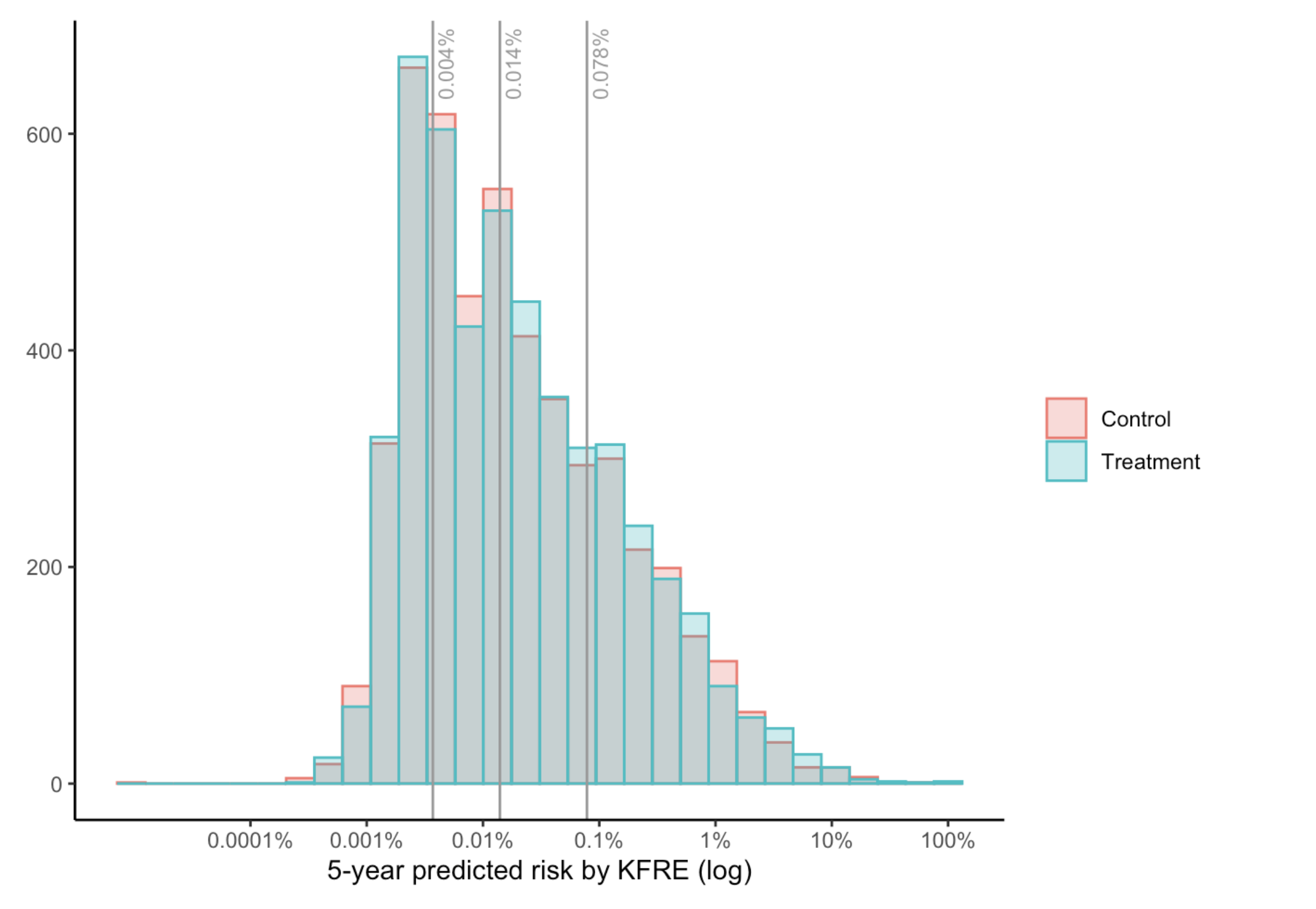

**Figure S2.** Baseline 5-year predicted kidney failure risk using the kidney failure risk equation among patients randomized to standard (red) and intensive (green) glycemic control. Vertical lines correspond to the 25^th^, 50^th^, and 75^th^ quantiles of the empirical distribution of 5-year kidney failure risk in the entire trial population based on the KFRE.

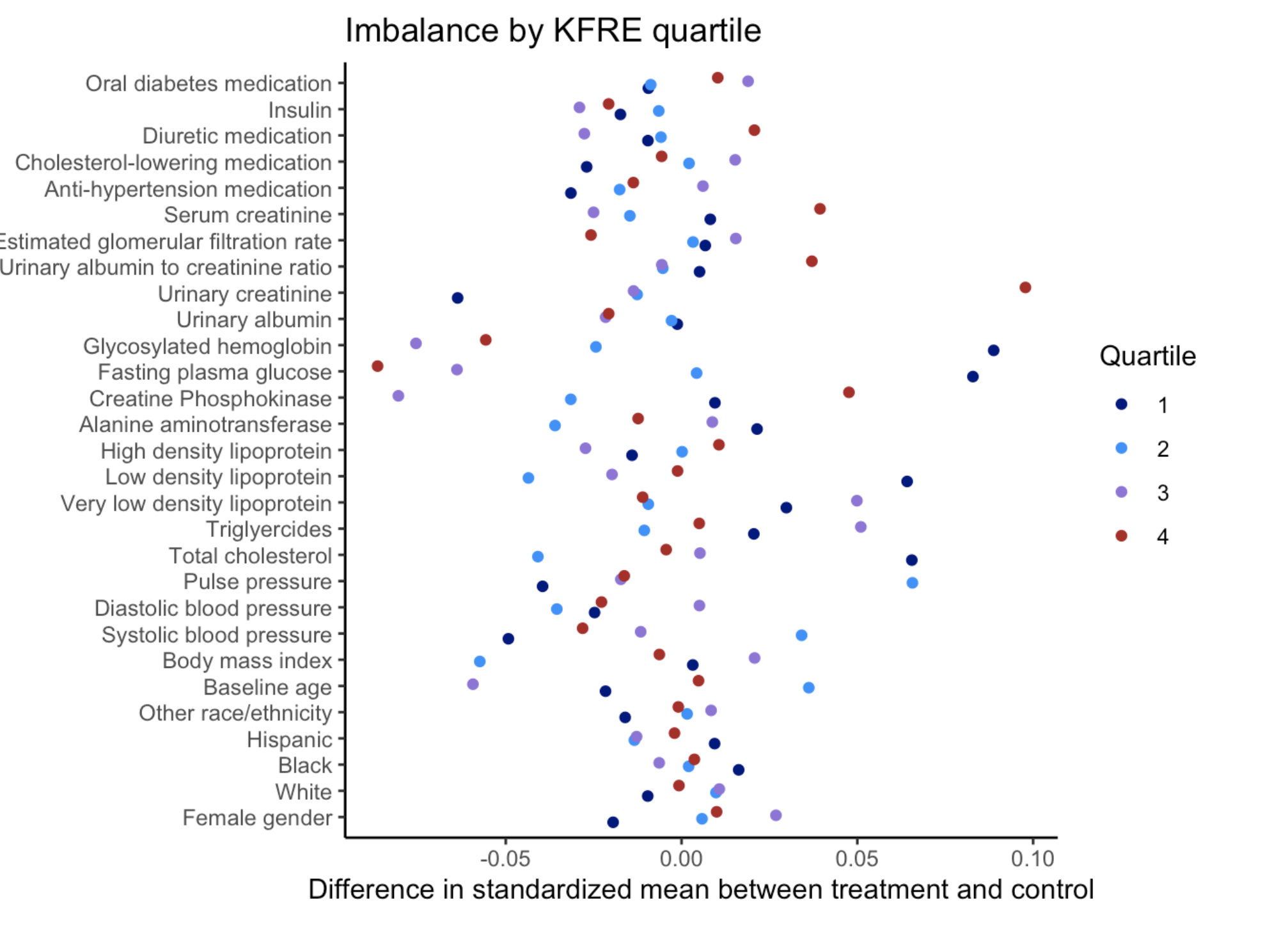

**Figure S3.** Covariate imbalance between treatment and control arms in ACCORD by quartile of KFRE. All standardized mean differences were within -0.10 to 0.10, indicating reasonable balance across baseline covariates. As such, additional adjustments were not performed when estimating the conditional average treatment effects within quartiles of predicted 5-year risk by KFRE.

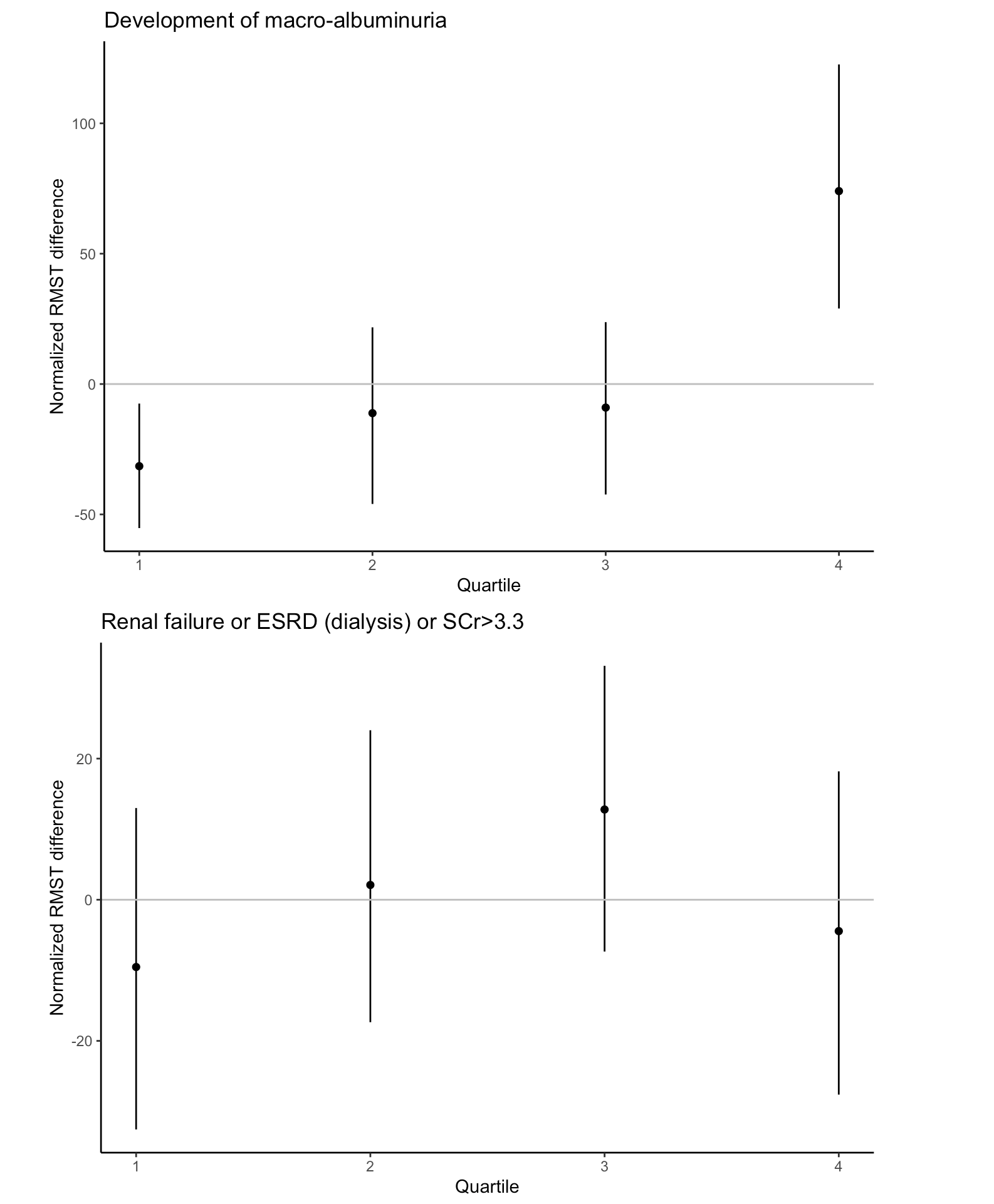

**Figure S4.** Heterogeneous treatment effects of intensive glycemic control on each renal microvascular outcome considered: (A) incidence macroalbuminuria and (B) renal failure. The x-axis displays each subgroup of patients, defined by quartiles of 5-year predicted risk by the KFRE. The y-axis displays the normalized restricted mean survival time (RMST) difference in days, defined as the RMST difference in the subgroup of interest minus the RMST difference in the entire trial. Normalized RMST values of zero indicate that the treatment effect in the subgroup of interest is equivalent to that in the entire trial population (no evidence of heterogeneous treatment effects). Normalized RMST values above zero (including the 95% confidence interval) indicate that the treatment effect in the subgroup of interest is larger than the treatment effect in the entire trial population (more beneficial); values below zero indicate that the treatment effect in the subgroup of interest is below that in the entire trial population (more harmful).

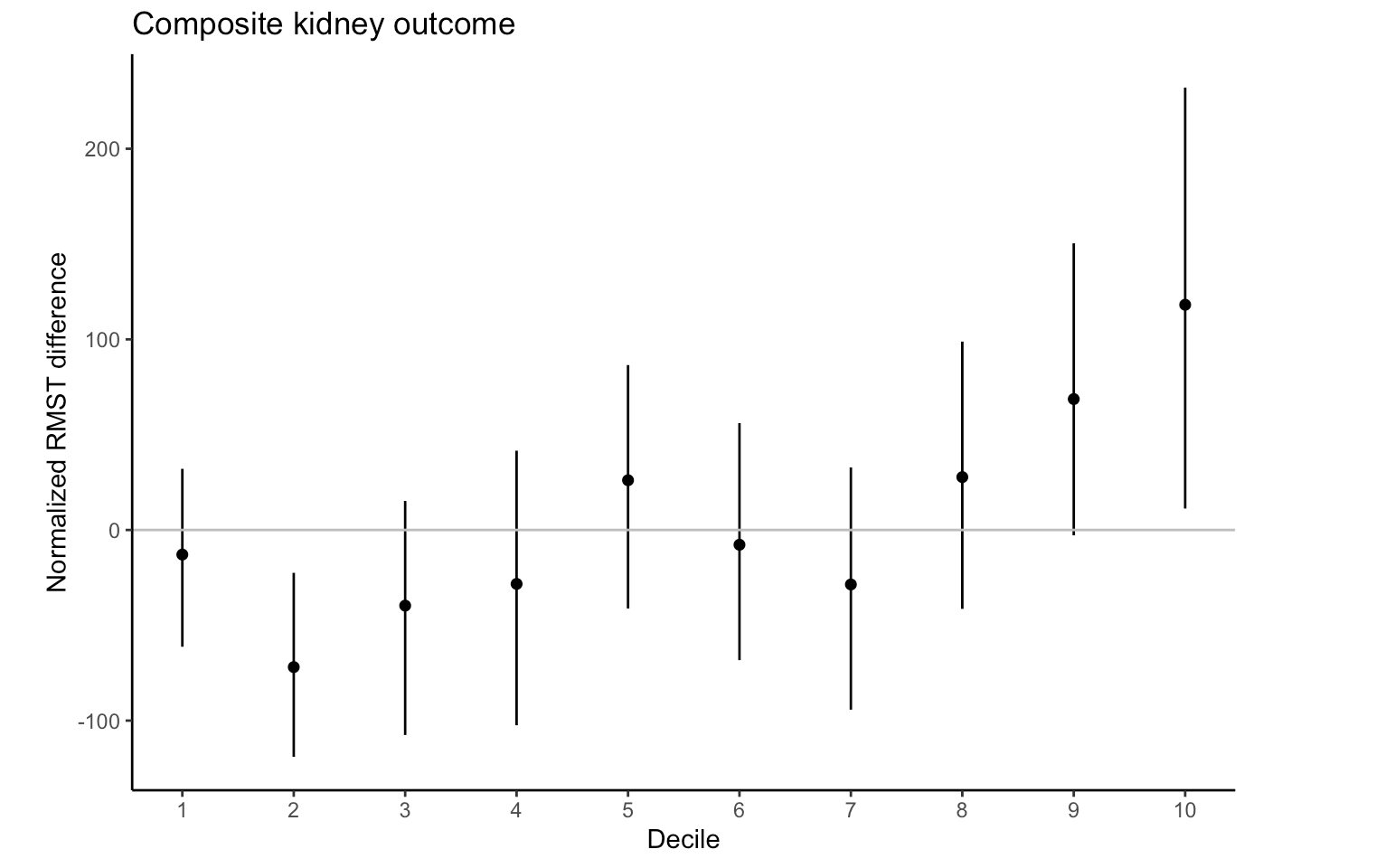

**Figure S5.** Heterogeneous treatment effects of intensive glycemic control on the composite kidney outcome within deciles of predicted 5-year kidney failure risk. The x-axis displays each subgroup of patients, defined by deciles of 5-year predicted risk by the KFRE. The y-axis displays the normalized restricted mean survival time (RMST) difference in days, defined as the RMST difference in the subgroup of interest minus the RMST difference in the entire trial. Normalized RMST values of zero indicate that the treatment effect in the subgroup of interest is equivalent to that in the entire trial population (no evidence of heterogeneous treatment effects). Normalized RMST values above zero (including the 95% confidence interval) indicate that the treatment effect in the subgroup of interest is larger than the treatment effect in the entire trial population (more beneficial); values below zero indicate that the treatment effect in the subgroup of interest is below that in the entire trial population (more harmful).

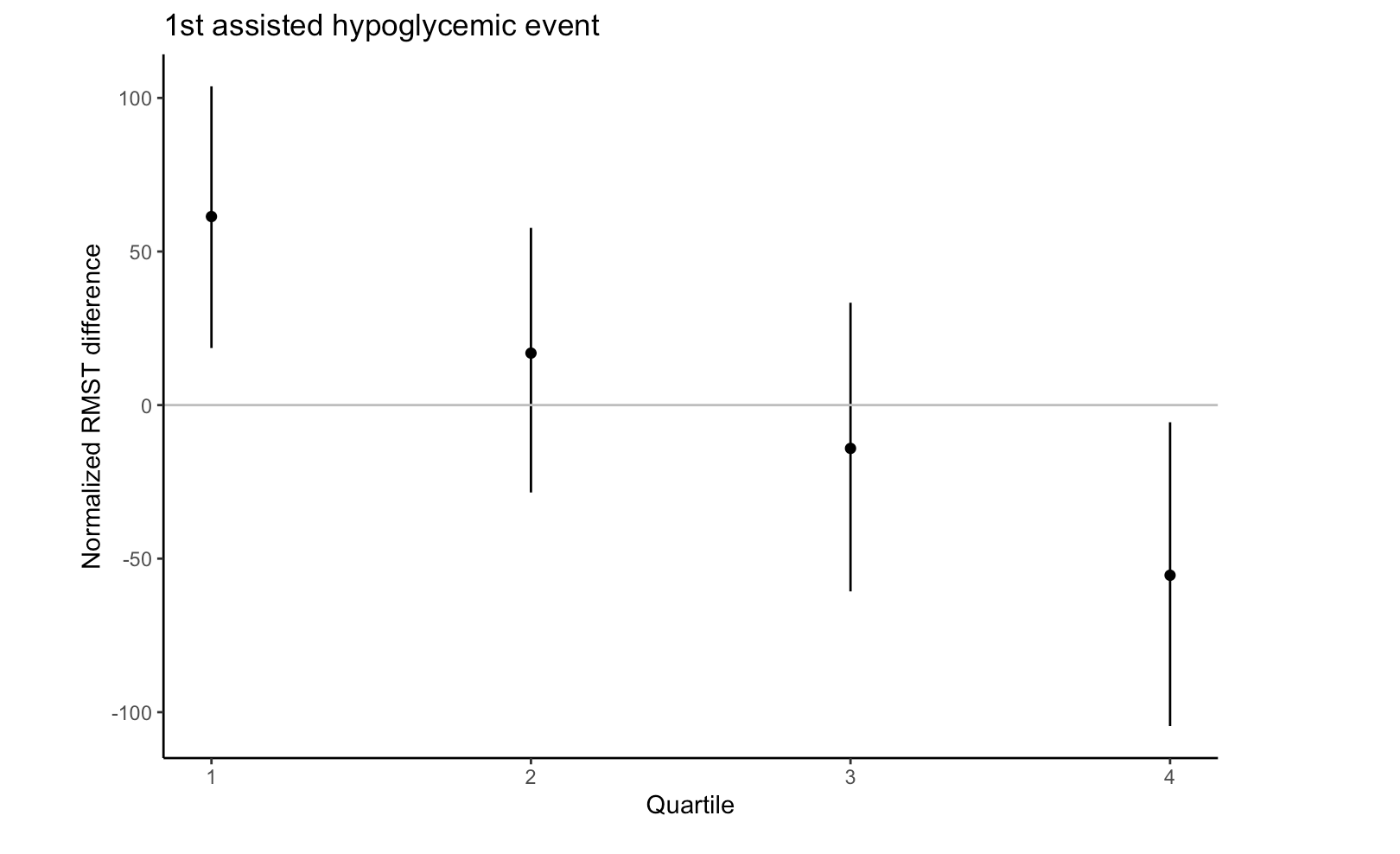

**Figure S6.** Heterogeneous treatment effects of intensive glycemic control on 1^st^ assisted hypoglycemic event by quartile of 5-year predicted kidney failure risk at baseline by KFRE. The x-axis displays each subgroup of patients, defined by quartiles of 5-year predicted risk by the KFRE. The y-axis displays the normalized restricted mean survival time (RMST) difference in days, defined as the RMST difference in the subgroup of interest minus the RMST difference in the entire trial. Normalized RMST values of zero indicate that the treatment effect in the subgroup of interest is equivalent to that in the entire trial population (no evidence of heterogeneous treatment effects). Normalized RMST values above zero (including the 95% confidence interval) indicate that the treatment effect in the subgroup of interest is larger than the treatment effect in the entire trial population (more beneficial); values below zero indicate that the treatment effect in the subgroup of interest is below that in the entire trial population (more harmful).

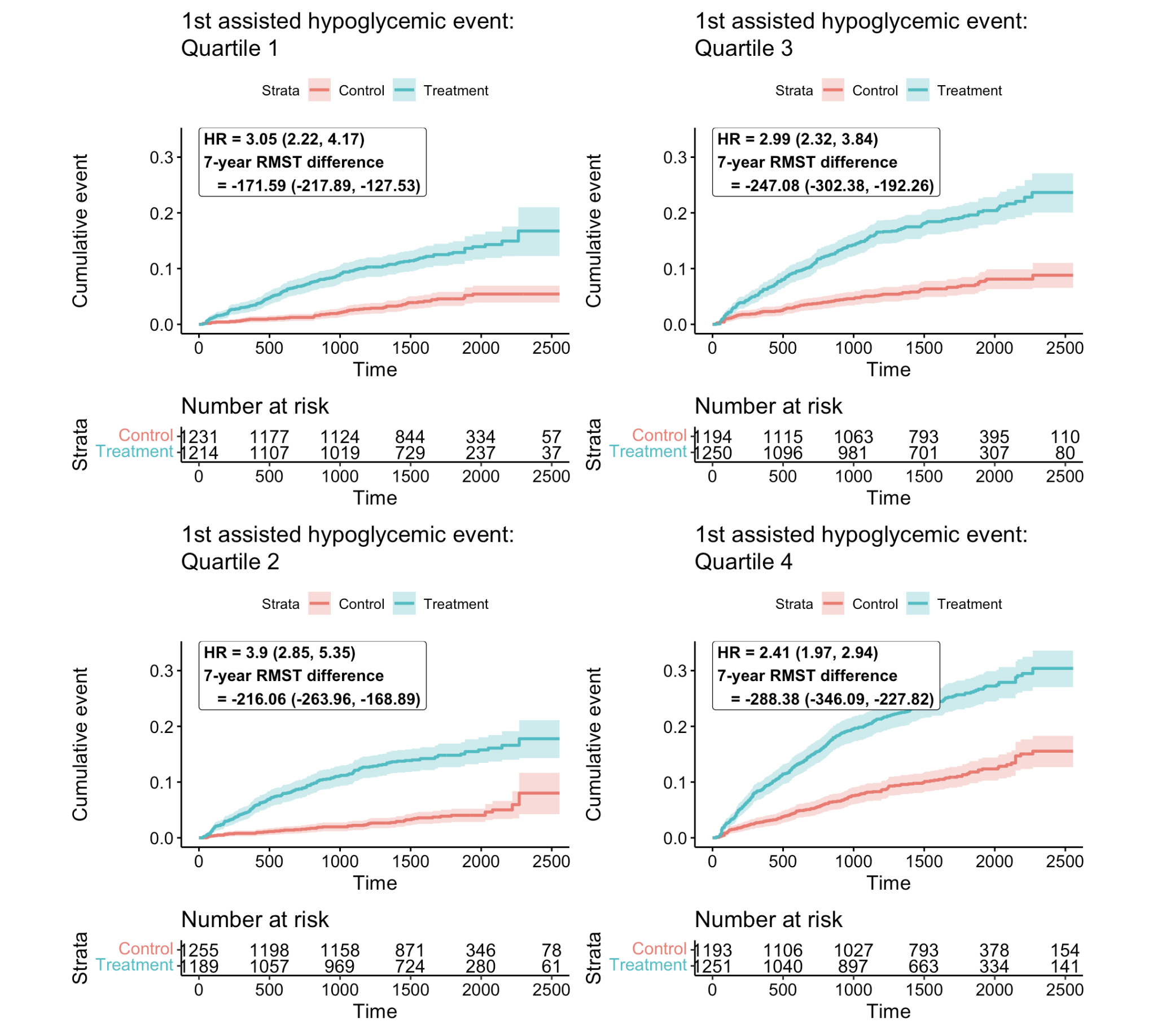

**Figure S7.** Heterogeneous treatment effects of intensive glycemic control on 1^st^ assisted hypoglycemic event by quartile of 5-year predicted kidney failure risk at baseline by KFRE. 7-year RMST differences are presented in days. Negative values indicate that intensive glycemic control shortened the time to the 1^st^ assisted hypoglycemic event.

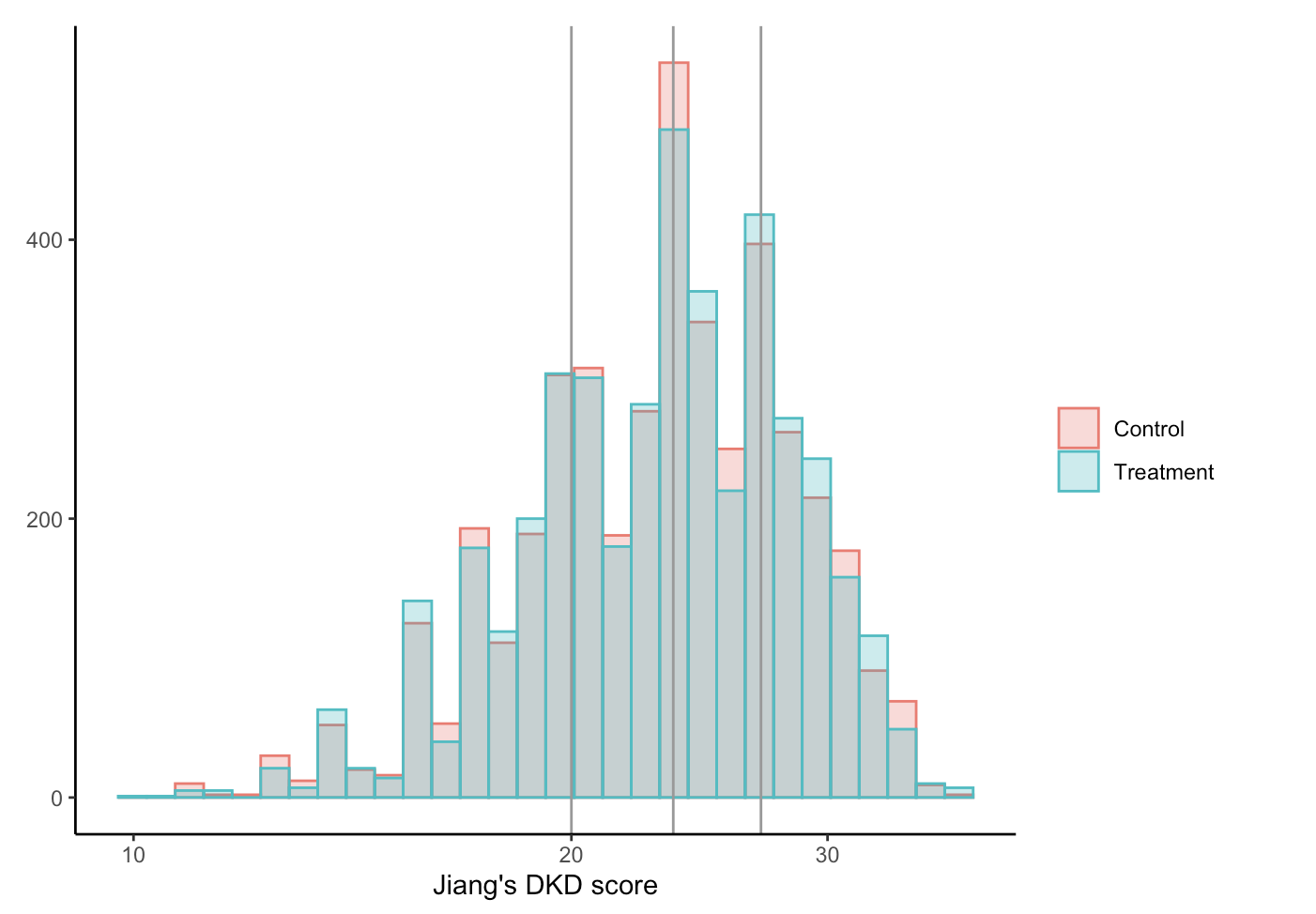

**Figure S8.** We explore whether Jiang et al.’s risk prediction model for early diabetic kidney disease (DKD) can be used to identify heterogenous treatment effects of intensive glycemic control on renal microvascular outcomes in ACCORD. The DKD risk score is based on nine risk factors: age, BMI, amoking status, diabetic retinopathy, HgbA1C, systolic blood pressure, HDL-C, triglycerides and UACR. The distribution of the DKD score by Jiang et al. in ACCORD. Vertical lines demarcate the 25^th^, 50^th^, and 75^th^ percentiles of the distribution.

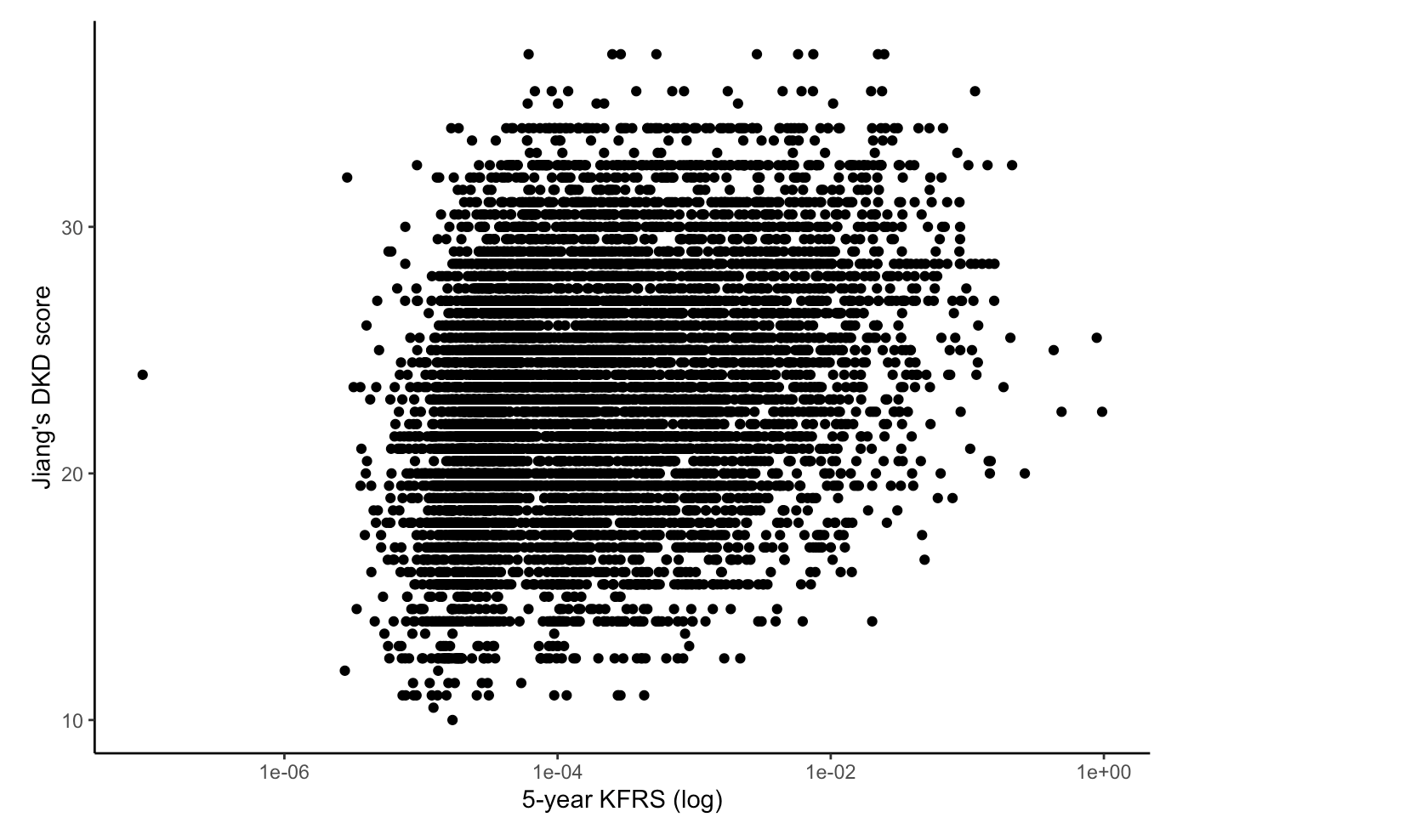

**Figure S9.** We explore whether Jiang et al.’s risk prediction model for early diabetic kidney disease (DKD) can be used to identify heterogenous treatment effects of intensive glycemic control on renal microvascular outcomes in ACCORD. The DKD risk score is based on nine risk factors: age, BMI, amoking status, diabetic retinopathy, HgbA1C, systolic blood pressure, HDL-C, triglycerides and UACR. Pairwise correlation between the 5-year kidney failure risk equation prediction and Jiang’s DKD score. Spearman’s rank correlation coefficient is 0.29.

**Figure S10.** Heterogenous treatment effects of intensive glycemic control within quartiles defined by Jiang et al.’s DKD risk score on (A) the composite kidney outcome, (B) incident macroalbuminuria, (C) renal failure of (D) all-cause death.

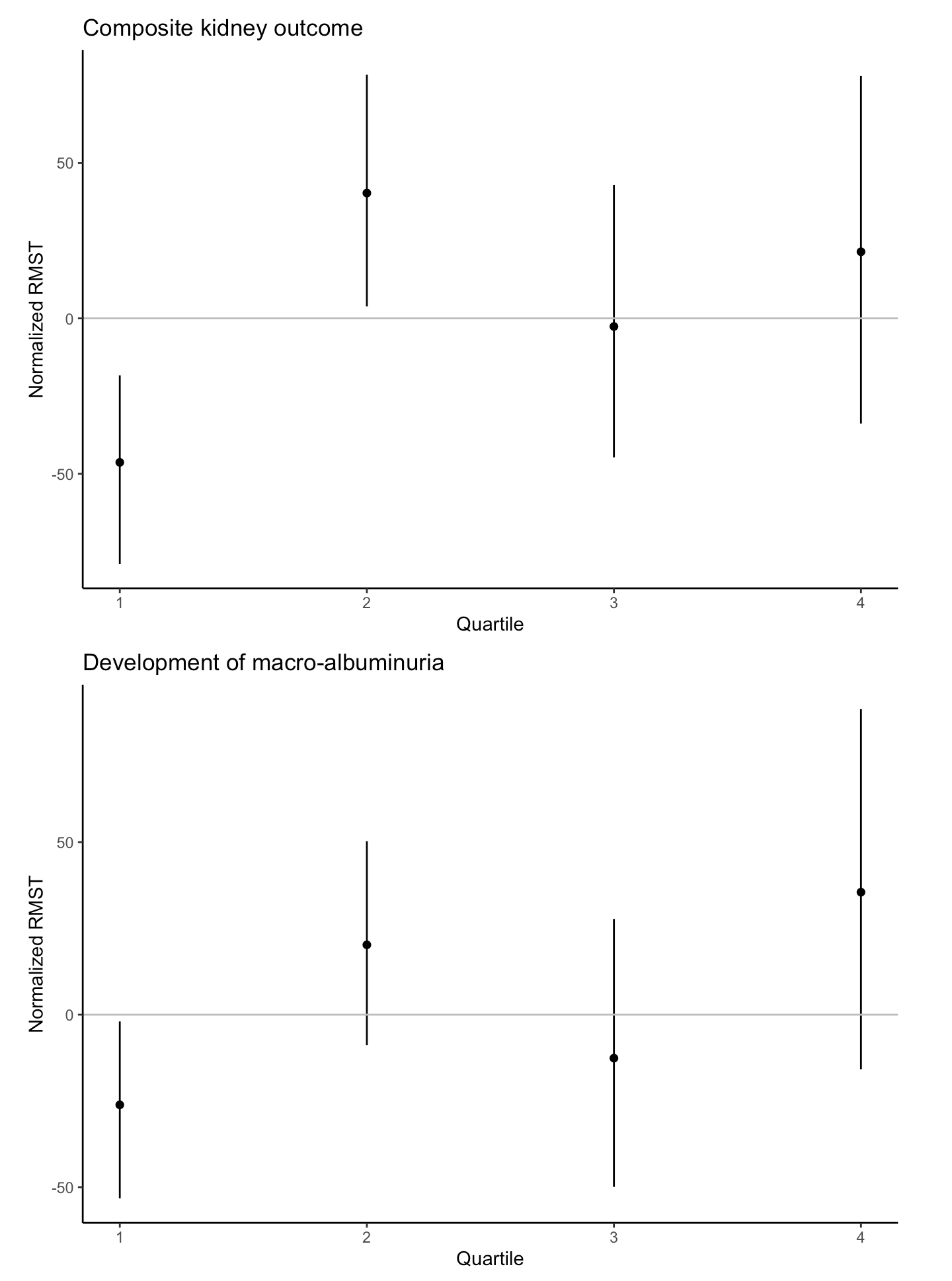

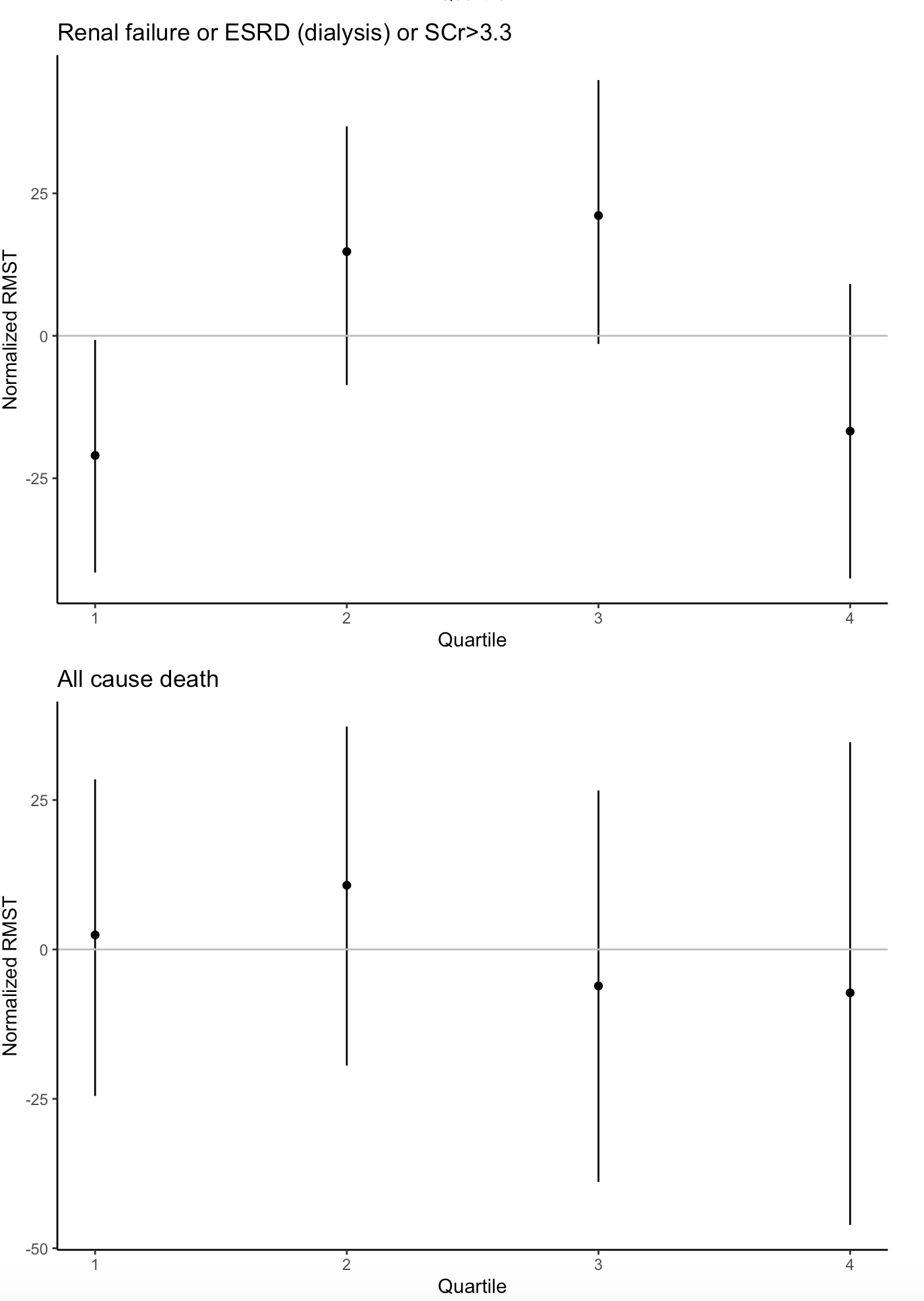
